# Detection of the SARS-CoV-2 spike protein in saliva with Shrinky-Dink© electrodes

**DOI:** 10.1101/2020.11.14.20231811

**Authors:** Julia A. Zakashansky, Amanda H. Imamura, Darwin F. Salgado, Heather C. Romero Mercieca, Raphael F.L. Aguas, Angelou M. Lao, Joseph Pariser, Netzahualcóyotl Arroyo-Currás, Michelle Khine

## Abstract

Using the children’s toy, Shrinky-Dink ©, we present an aptamer-based electrochemical (E-AB) assay that recognizes the spike protein of SARS-CoV-2 in saliva for viral infection detection. The low-cost electrodes are implementable at population scale and demonstrate detection down to 0.1 fg mL^−1^ of the S1 subunit of the spike protein.

## Introduction

In 2020, the World Health Organization (WHO) announced the respiratory disease COVID-19 outbreak to be a pandemic.^1^ The first cases of COVID-19 were in the Chinese city of Wuhan and it quickly spread to other continents. The high infection rate of the pathogen, severe acute respiratory syndrome coronavirus-2 (SARS-CoV-2), makes the management of the disease more difficult than previous coronaviruses, such as SARS-CoV-1 and MERS-CoV. Human-to-human transmission of SARS-CoV-2 can occur through droplets like other coronaviruses; however, there is a critical difference in SARS-CoV-2 viral load distribution throughout the time of infection that contributes to its rapid spread. Unlike SARS-CoV-1 or the common flu, whose viral loads are associated with the symptom onset, the high shedding of SARS-CoV-2 even among pre-symptomatic patients increases the risk of transmission of the virus. Several studies on the transmission of COVID-19 by asymptomatic patients have already been reported in recent literature.^2–4^ Due to the high transmission rate, measures such as “lockdowns” and “self-quarantining” were adopted, but the number of confirmed cases is still rising in many countries, including the United States. The widespread shortages of medical resources and materials reflect the overwhelming nature of the spread of this illness.

Due to the mode of transmission of COVID-19, the implementation of population-scale testing including asymptomatic people is recommended.^5^ Although the WHO recommends immediate diagnosis as a crucial management step to thwart the spread of infectious diseases, a massively deployable diagnostic for SARS-CoV-2 has yet to be realized.

In the U.S., 65.54 tests are performed per 1,000 people with a daily positive rate of 5.8 % as of October 25, 2020.^6,7^ The gold standard methods for diagnosing COVID-19 are based on polymerase chain reaction (PCR), including reverse transcription polymerase chain reaction (RT-PCR), which is a complex and expensive technique performed in centralized clinical laboratories. The turnaround time can be days, if not weeks, due to the currently overwhelming demand placed on laboratory equipment and manpower necessary to analyse specimens. Additionally, a major concern regarding the PCR-based tests is the high rate of false-negatives.^8^ It was shown that performing a RT-PCR test during the early days of infection (<4 days) can result in false-negative with rates well above 60% until after the fourth day of infection. The probability of false-negative is minimized to 20% on day 8, 3 days after the onset of the symptoms.^9^ In the days immediately after the exposure, tests based on RT-PCR add little diagnostic value for COVID-19, which is also the case for other infectious diseases such as HIV and hepatitis C.^9^ Another concern of current tests is the invasive sampling method. Nasopharyngeal swabs induce coughing and occasionally cause bleeding, which is a risk of transmission to healthcare workers. An easier and more accessible fluid is saliva. SARS-CoV-2 RNA was detected in saliva samples with concentrations ranging from 9.9 × 10^2^ to 1.2 × 10^8^ copies per mL.^10^ Non-invasive, painless saliva sampling decreases the risk of healthcare workers getting infected and allows specimen collection at the point of need, which can dramatically shorten waiting times for results. Using saliva not only removes the discomfort and increases the accessibility of current tests, but could provide results when nasopharyngeal swabs cannot.^11^ Yet, there still has not been (at the time of completion of this manuscript) a single screening or diagnostic test approved for over-the-counter (OTC) use by the FDA, which would allow users to collect their own saliva specimens, without having to mail them to a centralized laboratory for processing, and read results on the spot.

One alternative diagnostic approach is immunoassays, which can be used for self-testing of seroconversion status. Immunoassays detect the antibodies against SARS-CoV-2 and are more suitable to establish whether an individual was previously infected with COVID-19 because the antibodies persist in the body even after the infection. However, the immune system cannot immediately recognize and produce antibodies against a novel antigen, thus suffering from a period during which the virus can freely replicate. Therefore, detection of antibodies against SARS-CoV-2, or any other viral infection, is not recommended as a diagnostic tool or for contact tracing the spread of the disease.^12^ In contrast, during the critical first few days post infection, the viral load peaks, and with no indication of infection, the asymptomatic individual can effectively spread the virus to many people. Recent studies indicate that approximately 20% of infected patients remain asymptomatic but are infectious.^12,13^

Achieving highly specific and sensitive SARS-CoV-2 virus detection depends on enhanced accessibility to the viral antigen used in the diagnostic assay. Because the genetic material of the virus is encapsulated, PCR-based detection approaches require lysing steps prior to the measurement of viral load. This requirement adds complexity and variability that is suboptimal for point-of-need diagnostics. However, the need for specimen processing prior to detection can be eliminated by detecting virus components from the outermost coat layer, such as the spike protein, providing an attractive alternative for simplified virus detection. We note that virus copies present in saliva range from 10^2^ - 10^11^ copies per mL throughout the duration of the infection.^11,14–18^ Because these numbers are low, it is critical to establish an adequately low limit of detection for the assay. Here, we demonstrate an innovative platform that uses nucleic acid aptamers to recognize viral spike protein and successfully achieves the measurement of viral load within clinically relevant ranges of viral copies.

Critical to the development of sensitive diagnostic assays, aptamers have a significant advantage: the kinetic parameters of aptamer-target binding can be easily tuned to invoke a specific and measurable response.^19^ By tagging aptamers with a redox indicator such as methylene blue (MB), the change in MB electron transfer rate upon binding to the target molecule can be measured. Specifically, electrochemical aptamer-based (E-AB) sensors use this strategy to achieve response times on the order of seconds or faster and provide equilibrium results within 30 minutes.^20^ The limit of detection (LOD) for E-AB virus sensors depends on the target analyte, aptamer and most importantly, sensor design; the LOD can be brought to lower ranges through amplification methods to accommodate the low concentrations (aM - nM) of analyte present in physiological conditions.^21–23^ In 2009, an E-AB was used to detect the nucleocapsid protein of the SARS-CoV virus with a LOD of 2 pg mL^−1^, which falls in the lower range of concentrations of detectable viral load in nasopharyngeal and saliva samples.^24,25^ Most recently, a study was published in which an oligonucleotide targeting four different regions within the N-gene of the SARS-CoV-2 virus was used to detect SARS-CoV-2 viral RNA.^26^ The N-gene of the SARS-CoV-2 virus is located inside the virus and is released upon infection of cells. By contrast, each virus is thought to contain up to 100 spike proteins, based on the evaluation of genetically similar SARS-CoV viruses, each with a RBD capable of binding to an oligonucleotide specific to that region.^5,27–29^ For this reason, we chose the S1 subunit of the spike protein, which contains the RBD, as the detection target.

Recently, Song et al. discovered two aptamers targeting the RBD from SARS-CoV-2 by using an ACE2 receptor competition-based selection strategy (SELEX).^30^ For the biorecognition of our sensor, we selected the harpin-structured 51-base aptamer called CoV2-RBD-1C with a dissociation constant of 5.8 nM, which is comparable to that of existing antibodies raised against the spike protein. The measurements performed by Song et. al suggest that the aptamer binds to several amino acids of the RBD, which is essential for highly specific detection.^30^ Since the current diagnostic methods for COVID-19 are not capable to test in mass due to shortage of chemicals and invasive sampling method, we propose here a point-of-need E-AB platform based on Shrinky-Dink© electrodes, which are broadly available and manufactured at large scales, and an aptamer proven to be specific for the RBD region, to enable non-invasive detection of SARS-CoV-2 spike protein in saliva.

## Results and Discussion

We have previously demonstrated signal enhancement in the form of greater dynamic range of detection on electrodes made using the ‘80’s children’s toy, Shrinky-Dink ©, which saturates the color of a drawing made on a thermoplastic polystyrene sheet that shrinks when heated (Figure 1).^31,32^ Our group published on Shrinky-Dink Microfluidics in this journal in 2008 and it has since become a workhorse in laboratories around the world.^33,34^ In this study, we demonstrate using Shrinky-Dink wrinkled (SDW) electrodes for sensitive detection of the S1 subunit of the spike protein of SARS-CoV-2 with the purpose of providing a low-cost screening and diagnostic device for the COVID-19 infection. Figure 1 demonstrates the fabrication process of SDW electrodes, beginning with sputtering pre-stressed polystyrene with gold, followed by shrinking and immobilization of aptamers and 6-mercaptohexanol (MCH). The structured surface of the SDW gold electrode, shown in the scanning electron microscopy image, supports a high surface area for loading of MB-modified aptamers, which respond to the S1 protein domain by decreasing electron transfer currents from MB. The wrinkled gold surface has been extensively investigated in previous publications from our group and others.^31,32,35–39^

**Figure 1.**
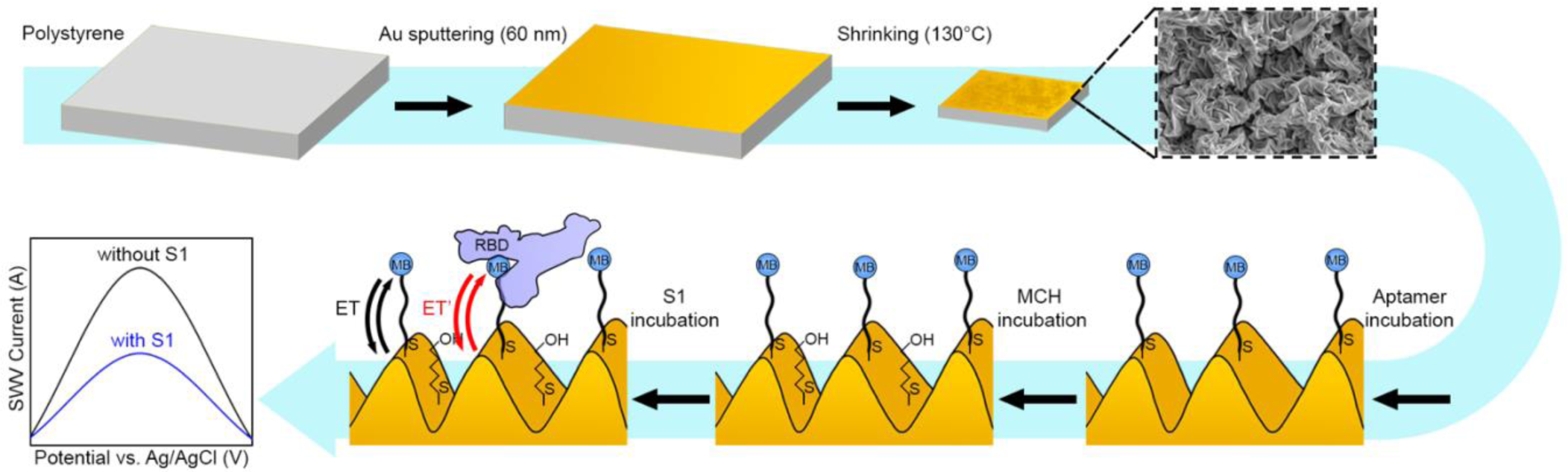
Process flow to create SDW E-AB sensors. First, a thin layer of gold is sputtered onto polystyrene plastic, which is shrunk to create wrinkles (SEM inset shows representative wrinkle morphology). The wrinkled surface is incubated with aptamers conjugated with MB, then incubated with MCH as the blocking molecule. After functionalization, the wrinkled surface was exposed to the S1 protein. Arrows indicate change in electron transfer with and without the spike protein attached (through the RBD). Graph illustrates change in current due to the change in electron transfer for spike bound MB on SDW electrodes upon addition of S1 protein.

To determine the functionality and affinity of the aptamers in the E-AB format, all initial experiments were first performed on commercial disc (CD) Au electrodes (CH Instruments, 2 mm diameter) in a standard 3-electrode electrochemical cell (Figure 2A). The commercial gold electrodes were functionalized with the aptamers and backfilled with blocking MCH monolayers. Upon exposure of the electrode surface to a buffered solution containing S1 protein, we observed a decrease in the MB peak height of square wave voltammograms (Figure 2B). The binding of a relatively large protein, such as the S1 protein (78.3 kDa), to the MB-modified aptamer produces an increase in resistance to the transfer of electrons from MB to the electrode (presumably via steric hindrance), leading to a decrease in the MB peak similar to that presented in the work of Kang et al.^40^ In the E-AB sensors, the mechanism of detection relies on binding-induced conformational changes that, in turn, alter the electron transfer rate between a redox reporter and the surface of the electrode.^41,42^ The distance between the reporter and the electrode surface directly affects electron transfer kinetics in the system; at small distances, and with little obstruction, electron transfer occurs at a faster rate than at greater distances or in the presence of obstructing molecules. These differences in electron transfer rates translate to differing time constants of current passed, which become relevant when considering measurement parameters.^43^ For the SARS-CoV-2 RBD aptamer used in this study, no conformational change has yet been demonstrated. Thus, we speculate that the changes in signal observed in this study upon target binding are due to the physical obstruction of electron transfer from the reporter to the gold surface by bound spike proteins.

**Figure 2.**
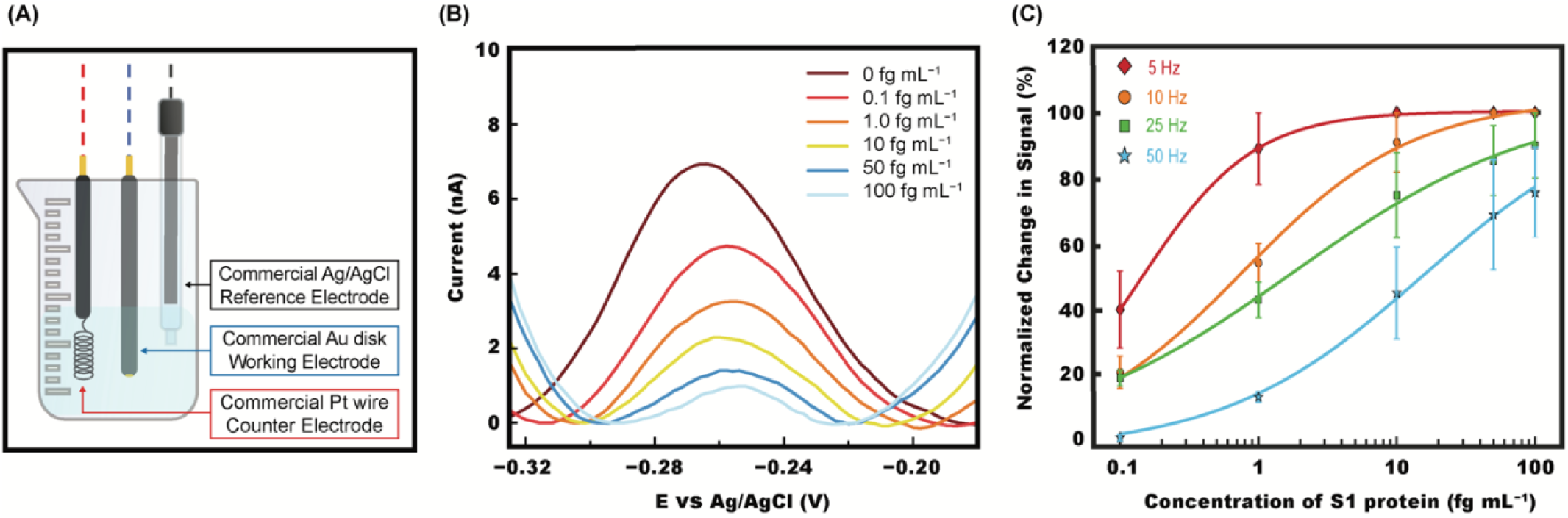
(A) Beaker cell configuration with CD electrode. (B) Raw peak height change in current with increasing concentrations of S1 protein in phosphate buffer solution on CD electrodes. (C) Titration curves collected at various frequencies.

The change in peak current correlates with increasing concentrations of S1 protein, as shown in Figure 2B. We evaluated the change in signal in response to increasing S1 protein concentrations on CD Au electrodes by performing a titration curve in phosphate-buffered solution (Figure 2C). Measurements were taken at square-wave frequencies ranging from 5 to 50 Hz. Non-linear regression using the Hill isotherm was used to determine the optimal square-wave frequency producing maximum signal gain and broadest dynamic range. This analysis resulted in Hill parameters that reported varying receptor-ligand affinity (*K*_*D*_) and binding stoichiometry with square-wave frequency. The Hill coefficient *n* was ∼ 1 at 5 Hz and the data displayed highest sigmoidicity at this frequency, indicating single binding site physics for the interaction between the S1 protein and the aptamer. Frequencies above 5 Hz produced isotherms with slightly lower n values (e.g., n = 0.78 at 50 Hz) but broader dynamic range. Based on these calibrations, 10 Hz was chosen as the optimal compromise between sensor affinity and sensitivity for our platform.

The signal change in response to addition of S1 protein on CD electrodes confirmed the viability of the chosen aptamer probes; however, using CD electrodes for screening and diagnostic applications is not feasible. CD electrodes cost $90 each, and given the contagious nature of the disease, reusing them presents significant practical challenges. For a truly deployable and scalable approach, the entire sensor must be disposable. With such considerations of cost of production and scaling in mind, we investigated electrodes fabricated with a simple sputtering deposition process on a commercial polystyrene substrate. Before the shrinking process, the adhesion of sputtered gold on polystyrene is too weak and the gold thin film can delaminate. When shrunk, however, the resulting SDW electrodes are extremely robust and retain their original surface area, resulting in enhanced current density and dynamic range.^44^ Because of the advantages in cost and performance, we initiated experiments on SDW gold electrodes (Figure 3A) with the goal of detecting the S1 protein directly in saliva.

**Figure 3.**
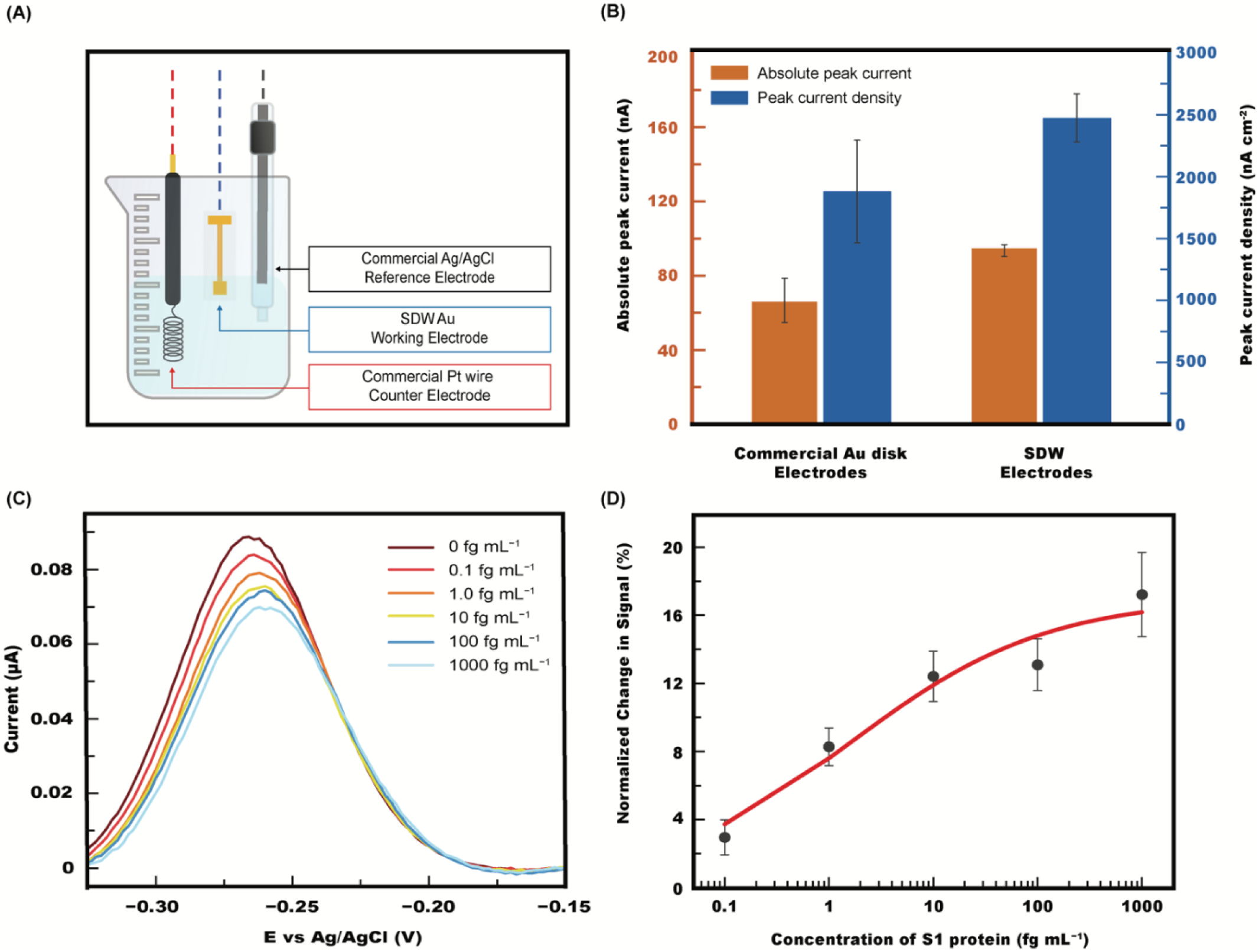
(A) Beaker cell configuration with SWD electrode. (B) Raw peak height change in current with increasing concentrations of S1 protein in 10% saliva on SDW electrodes. (C) Normalized change in signal produced from sequential incubations of saliva spiked with increasing concentrations of S1 protein minus signal from sequentially incubated blank saliva samples. Hill fit represented in red. (D) Methylene blue absolute peak current and peak current density comparison between CD and SDW electrodes with equivalent geometric areas.

The total accessible electrochemically active surface area (EASA) of SDW electrodes was confirmed to be greater than that of CD electrodes, as calculated by integration of the reduction peak of gold oxide in sulfuric acid for the respective electrode types (Figure S1). The electrodes were of equivalent geometric areas. The wrinkles provide a diffusion-based challenge to surface coverage, as previously described.^31^ With longer incubation time, however, this limitation was overcome. The outcome was a greater number of moles of aptamers tethered to the surface of SDW electrodes relative to the same geometric area of CD electrodes. The increased aptamer density resulted in greater absolute MB peak heights on SDW electrodes. Figure 3B demonstrates the MB peak current densities of CD and SDW electrodes are comparable, with the CD electrodes displaying greater variability as measured by the magnitude of the standard deviation between 3 independent electrodes.

To determine whether the introduction of S1 protein to saliva would produce a differentiable signal change relative to untreated saliva, titration curves were built by measuring square wave voltammograms at 10 Hz with SDW electrodes exposed to varying concentrations of S1 protein. The change in signal with increasing concentrations of S1 protein is shown in Figure 3C. A slight shift in the reduction voltage of MB was observed and attributed to binding-induced changes in the local pH at the surface of the electrode. The titration curve displayed in Figure 3D was derived from two electrode sets. One set was incubated in saliva, and the second in saliva spiked with S1 protein. Both sets were incubated in saliva for the same total amount of time. The two separate datasets can be found in Figure S2.

As evidenced from Figure 3D, SDW electrodes showed a smaller overall signal change than CD electrodes; we attribute this to the SDW electrode measurements being performed in saliva, as opposed to buffer solution as in Figure 2. A loss of signal gain in measurements collected from human media compared to buffer solution has been documented previously.^45^ The standard error of signal change was lower in SDW electrodes than in CD electrodes, however, which demonstrated the robustness of the fabrication process of SDW electrodes.

To improve the portability of our sensor, we next fabricated an entire miniaturized electrochemical cell, abbreviated as “mini cell”, using the same shrinking process used for individual SDW electrodes. The SDW mini cells’ working and counter electrodes were created by sputtering gold onto the pre-stressed polymer substrate and heated to induce shrinking. Ag/AgCl ink was applied as a reference electrode on the shrunk substrate; thus, all three electrodes were located on one substrate, allowing electrochemical measurements within a 250 µL droplet instead of the 20 mL used in the beaker cell. The performance of the SDW mini cell was evaluated by characterizing the gold oxidation and reduction in sulfuric acid. Compared to measurements collected using the commercial reference and counter electrodes, the SDW mini cell demonstrated similar performance, with slightly increased peak-to-peak separation (ΔEp) between oxidation and reduction peaks (Figure S3).

We observed that during the cleaning protocol performed in sulfuric acid, the area under the peak of the SDW mini cell increased more than that of individual SDW electrodes (Figure 4A). We hypothesize that because the SDW mini cell contains an additional source of gold (the counter electrode), there may be a greater rate of gold deposition onto the working electrode occurring during the sulfuric acid cycling, leading to a greater final EASA in the SDW mini cell. The peak height stabilized by the 120^th^ cycle, after which the functionalization of the working electrode on the mini cell proceeded in the same way as for SDW electrodes (Figure 4B). We next evaluated the SDW mini cell for detection of the S1 protein in saliva. The SDW mini cell was connected to the PalmSens USB drive-sized potentiostat (Figure 4B) with alligator clips to demonstrate collection of measurements in any location (non-lab environment).

**Figure 4.**
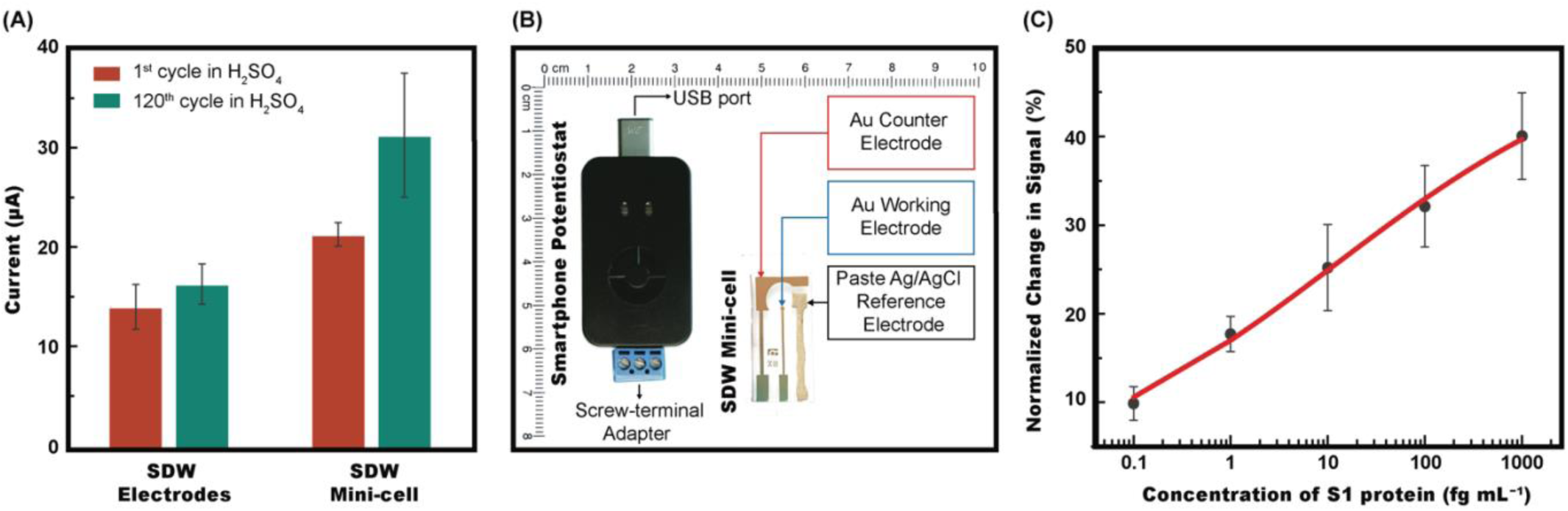
(A) Comparison of change in area under the gold reduction peak during cycling in sulfuric acid between individual SDW electrodes and SDW mini cells. Peak height and area under the peak stabilized by 120 cycles at 1000 mV s^−1^. (B) Sensit Smart (Palmsens) potentiostat next to a SDW mini cell with scale provided in centimeters. Electrode arrangement (left to right): counter (60 nm Au), working (60 nm Au) and reference (Ag/AgCl ink). (C) Titration curve of S1 protein performed at 10 Hz in 10% saliva. Signal change normalized to 10% saliva without S1 protein.

The signal gain of 40% at the highest concentration of spike protein was greater in the SDW mini cells than in the individual SDW electrodes, in which the maximum change was approximately 16% (Figure 4C). Because the aptamer functionalization protocol (incubation time, aptamer and MCH concentrations) was the same for both individual SDW electrodes and SDW mini cells, the larger surface area of the SDW mini cell working electrode resulted in a lower probe density. This was also reflected in the differences in peak current densities of MB between individual SDW electrodes and SDW mini cells. Previous studies have shown that a lesser probe density can contribute to greater sensitivity to signal change, which was found to be the case in this study.^46,47^ We therefore demonstrate the detection of the S1 protein in the same range tested for individual SDW electrodes, but with greater signal change in the SDW mini cells. From these results, we believe the SDW mini cell to be a promising option for further investigation of portable individual tests.

The SDW mini cell demonstrated detection of as little as 0.1 fg mL^−1^ S1 protein in 10% saliva, with the full comparison to system noise described in the SI. In comparison, a recent study utilizing the same aptamer to target the RBD region of the spike protein achieved a LOD six orders of magnitude greater in buffer solution than the lowest concentration presented here.^48^ Additionally, our low-complexity process does not involve the use of costly materials and does not rely on enzymatic reactions or biotinylation, which is known to cause interference with clinical samples.

## Conclusion

Although biosensors have largely utilized antibodies to recognize target molecules or pathogens in the previous decades, an equally specific alternative has recently emerged. Aptamers that selectively bind to specific targets have extensively been demonstrated to be promising as a diagnostic tool for disease detection through molecular recognition, and have been prominently investigated.^21,45,49–55^ In this study, an aptamer specific to the RBD region of the SARS-CoV-2 spike protein was selected as the probe for the detection of the SARS-CoV-2 in saliva.

The SDW electrodes used in this study demonstrated sensitive detection of the S1 protein at 0.1 fg mL^−1^ in pooled saliva diluted to 10%. The facile fabrication process and the sensitivity to low concentrations of S1 protein indicate a promising opportunity to increase the accessibility of SARS-CoV-2 screening and widespread testing of asymptomatic people. The biosensor still needs to be validated to detect the whole virus but given the location of the S1 protein at the outermost layer of the virus, we expect that this should not be an issue.^26^

In future studies, the varying performance at different frequencies as well as different probe densities should be investigated to better understand aptamer behaviour on the molecular level. Additionally, we expect using aptamers specifically designed for binding-induced conformational changes would produce even more sensitive results than with the aptamer used in this study, which has no confirmed conformational change upon binding with the S1 protein. Importantly, this SDW platform allows for ease in substituting different aptamers, enabling detection of a wide range of analytes for future epidemics.

## Materials & Methods

### Chemicals

DNA Aptamers were chosen according to prior literature.30 The oligonucleotide modified with methylene blue at the 5’ end and a thiol group at the 3’ end was purchased from Integrated DNA Technologies, Inc with the following sequence: /5MeBlN/CA GCA CCG ACC TTG TGC TTT GGG AGT GCT GGT CCA AGG GCG TTA ATG GAC A/3ThioMC3-D/.

Aptamer reducing buffer was purchased from Base Pair Biotechnologies. Aptamers were diluted in phosphate buffer solution. The S1 protein was purchased from GenScript (Cat. No. Z03501). 6-mercaptohexanol (MCH), monobasic potassium phosphate (KH_2_PO_4_) and dibasic potassium phosphate (K_2_HPO_4_) were purchased from Sigma Aldrich. Pooled saliva was purchased from CellSciences.

Phosphate buffer solution was prepared with KH_2_PO_4_ and K_2_HPO_4_ at 0.1 M. Sulfuric acid (H_2_SO_4_ - Sigma Aldrich) was prepared in water at 0.5 M. In experiments involving saliva, the saliva was diluted to a final concentration of 10% in phosphate buffer.

### SDW Electrode Fabrication

The electrode design was created by applying an adhesive polymer (Grafix Frisket Film, Grafix Arts, OH) shadow mask stencil to the polystyrene (Grafix Shrink Film KSF50-C, Grafix Arts, OH) prior to sputtering. A Q150R Plus – Rotary Pumped Coater was used to sputter 60 nm of gold onto the pre-stressed polystyrene to create the working electrode. The substrate was de-masked and placed into a conventional oven at 130°C for 13 minutes until fully shrunk. Following this step, the electrode was treated with oxygen plasma for 3 minutes to achieve temporary hydrophilicity to ensure full surface wettability during the following cleaning step.

### SDW mini cell Fabrication

The SDW mini cells’ working electrode and counter electrodes were created by applying an adhesive mask to the polystyrene prior to sputtering. Following the same protocol as for individual SDW electrodes, 60 nm of gold was sputtered onto the pre-stressed polystyrene to create the working and counter electrode. The substrate was then de-masked and placed into a conventional oven at 130°C for 13 minutes until fully shrunk. Following this step, the reference electrode was painted onto the shrunk polystyrene using commercially available Ag/AgCl ink (Creative Materials). The working and counter electrodes were treated with oxygen plasma for 3 minutes to achieve temporary hydrophilicity, while the Ag/AgCl electrode was covered to prevent oxidation.

### Cleaning electrodes

Commercial electrodes were polished using the 0.5 µm-sized Ag particles included in the kit provided with the electrodes prior to cycling in sulfuric acid. The electrodes were then individually immersed in a 0.5 M solution of H_2_SO_4_ and subjected to 120 cycles at 1000 mV s^−1^ followed by five cycles at 100 mV s^−1^ in the potential window of 0 – 1.5 V.

Individual SDW electrodes were similarly immersed in a 0.5 M solution of H_2_SO_4_ and subjected to 120 cycles at 1000 mV s^−1^ followed by five cycles at 100 mV s^−1^ in the potential window of 0 – 1.5 V.

SDW mini cell electrodes were cleaned by drop-casting a 250 µL drop of 0.5 M solution of H_2_SO_4_ onto the surface, ensuring coverage of all three electrodes’ working areas, and subjected to 120 cycles at 1000 mV s^−1^ followed by five cycles at 100 mV s^−1^ in the potential window of 0 – 1.5 V.

The last cycle at 1000 mV s^−1^ for each electrode type is shown in Figure S3.

### Aptamer preparation

Aptamers were received in liquid form and diluted to a concentration of 100 µM. For each experiment, a small volume of aptamer solution (100 µM) was combined with a reducing buffer (Basepair Technologies) at a 1:2 volume ratio for one hour to reduce the 3’ ends of the aptamers. Following the reduction step, the solution was diluted with phosphate buffer solution to a final concentration of 1 µM. 1 µM is an excessive concentration to use, but additional studies must be carried out to determine optimal concentration of probes for incubation. Optimal probe density must also be investigated in order to confirm optimal sensor sensitivity. Probe density affects the magnitude of signal change and is assumed to be a function not only of initial concentration of applied aptamers, but also of incubation time and temperature.^56^ In the case of oligonucleotides greater than 24 bases, such as in this investigation, the probe density is greatly affected by incubation time.^46^ From the SDW mini cell experiments, it was evident that a lesser probe density contributed to enhanced signal gain.

### Functionalization of electrodes with aptamers

A 30 µL of the aptamer solution at 1 µM was applied to each electrode and allowed to incubate 4 and 8 hours, respectively, for CD and SDW electrodes, at room temperature. The aptamer incubation time was optimized for SDW electrodes; from previous investigations, it is clear that the wrinkled morphology presents diffusion limitations for molecular access to the surface.^31^ Thus, the incubation time of aptamers was increased to ensure adequate surface coverage by the aptamer probes. During this time, the aptamers formed thiol bonds with the gold surface. The presence of the characteristic MB peak at –0.28 V confirmed the attachment of the aptamer to the gold surface. The peak height collected from each type of electrode (CD, SDW individual and mini cell) was normalized to their respective EASA for comparison of current density. The current density was assumed to correlate with the density of MB and the aptamer probes to which it is attached on the electrode surface. The results of a two-sample t-test show a rejection of the null hypothesis at the 5% significance level that the current densities of CD electrodes and individual SDW electrodes have an equal mean, indicating that the individual SDW electrodes may have a greater probe density. We believe this may be attributed to the longer incubation time chosen for SDW electrodes to compensate for diffusion limitations posed by wrinkles. In contrast, due to the greater EASA of the SDW mini cell working electrodes, the probe density was found to be lesser than that of individual SDW electrodes.

### Blocking of electrodes with MCH

Blocking the electrode surface avoids nonspecific binding with interfering species in real samples, enhances the stability of aptamers, and passivates any remaining EASA. A six-carbon blocking molecule, MCH, was chosen as the blocking agent because it has been shown to create more stable monolayers than a 2- or 3-carbon molecule, although at the expense of greater conductivity of the SAM achieved using shorter chains.^46^ After the incubation with the aptamers, the electrodes were rinsed and incubated with 30 µL of 3 mM MCH in phosphate buffer for 18 and 39 hours, respectively, for CD and SDW electrodes (both individual and mini cell), at 4°C.

### Titration curve of S1 protein on CD electrodes

After functionalization and blocking, the commercial gold electrodes were measured in 20 mL of phosphate buffer with a Gamry Reference 600 potentiostat with a commercial Ag/AgCl reference electrode (Gamry) and a platinum coil (BASi) as the counter electrode. Following the collection of a baseline measurement, the electrodes were incubated for one hour with increasing concentrations of S1 protein in phosphate buffer solution.

### Titration curve of S1 protein on SDW gold electrodes

Six SDW gold electrodes were incubated with aptamers and blocked with MCH, as described above. All six electrodes’ MB peak heights were measured after every functionalization step in phosphate buffer solution with the same setup for commercial electrodes. Three of the electrodes were incubated for one hour with 10% pooled saliva in phosphate buffer then rinsed carefully with phosphate buffer solution. The three remaining electrodes were incubated for one hour with 10% pooled saliva containing 0.1 fg mL^−1^ S1 protein. The MB signal from all six electrodes was measured again in phosphate buffer solution. Subsequent incubation steps were performed in the same way, with all electrodes incubated with either un-spiked saliva containing increasing concentrations of S1 protein with every incubation. All electrodes were rinsed before all measurements. Measurements were collected after establishing signal stability across 10 consecutive measurements in phosphate buffer solution at 10 Hz (Figure S4) to ensure signal stability was within 5% normalized change.

### Titration curve of S1 protein on SDW mini cell

The working electrodes of the SDW mini cells were incubated with 10% saliva for 1 hour containing no S1 protein to constitute the zero point of the titration curve. The SDW mini cells were then incubated with 10% saliva containing the following concentrations of S1: 0.1, 1.0, 10, 100, and 1000 fg mL^−1^. The SDW mini cell was rinsed following each incubation period at 1 hour, and a 250 µL drop-cast of phosphate buffered solution was applied to cover all three electrodes to collect measurements from the PalmSens Sensit Smart.

### Electrochemical measurements

Gamry Reference 600 potentiostat. Conventional potentiostat settings were measured in a phosphate buffer solution with a potential window starting from −0.35 to −0.15 V. Frequencies measured at: 5, 10, 25, 50, 75, 100, 150, 200 and 250 Hz. The amplitude was set to 25 mV with a step size of 2 mV.

PalmSens Sensit Smart. All mini cell measurements were measured in a phosphate buffer solution with a potential window starting from −0.7 to −0.2 V. Frequencies measured at: 5, 10, 25, 50, 75, 100, 150, 200 and 250 Hz. The amplitude was set to 25 mV with a step size of 2 mV.

## Data Availability

All data is contained within the manuscript and supplemental information document.

## Acknowledgements

We would like to thank Hui Li for graciously providing guidance on signal stability. We thank Julien Goavec for the initiation of mini cell experimentation. HCRM gratefully acknowledges the support provided by the National Institute of Health through the NIH-MARC U-STAR training grant T34GM136498. AHI acknowledges the financial support from grant #2017/05362-9, São Paulo Research Foundation (FAPESP).

