## Supplemental Information for "Detection of the SARS-CoV-2 spike protein in saliva with Shrinky-Dink© electrodes"

***MB peak density calculation:***

The CD and SDW electrodes' electrochemically active surface area (EASA) was investigated using the area under the peak of their respective voltammograms collected in 0.5 M sulfuric acid. A representative integrated peak from a CD electrode is shown in Figure S1. The preparation of electrodes included the performing 30 cycles of cyclic voltammetry in 0.5 M sulfuric acid at 1000 mV/s followed by 5 additional cycles at 100 mV/s. In our previous work, it was determined that integration of the peak of the voltammogram taken at a scan rate of 1000 mV/s produced the most accurate value for presumed EASA. Thus, the EASA of electrodes was determined by dividing the charge of the electrode by the characteristic surface charge of gold (390 uC/cm$^2$). The average peak current of MB measured in PBS was then divided by the calculated EASA to compare current density between standard and wrinkled electrodes.

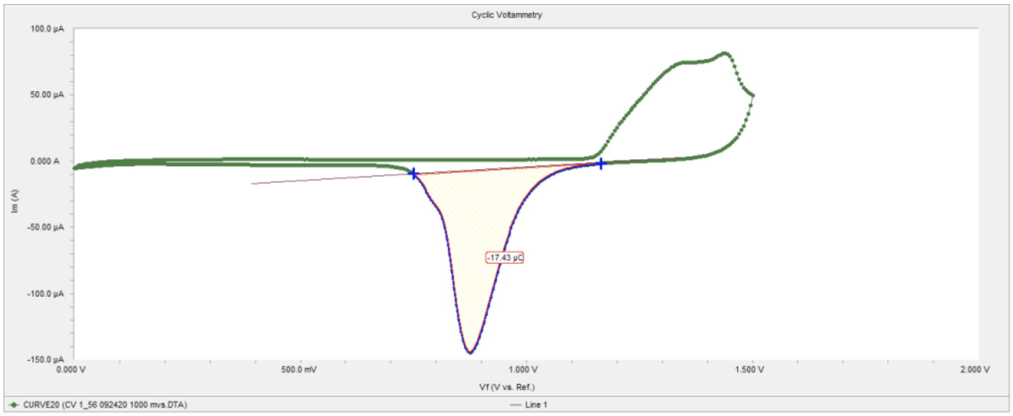


**Figure S1.** Wrinkled electrode reduction peak collected in 0.5 M sulfuric acid, from which charge was determined for EASA calculation.

Compared to a standard commercial electrode, the available surface area for binding aptamers is greater per geometric area. This translates to a greater total amount of probes capable of recognizing the S1 protein, which we hypothesize to be the cause of the greater dynamic range of detection. This theory is supported by our comparison of MB peak heights between standard and wrinkled electrodes.

The comparison of MB peak heights collected from 3 standard electrodes and 3 wrinkled electrodes shows a slightly greater absolute current on wrinkled electrodes (Figure 3D). We conclude that there is a greater amount of MB present on the wrinkled surface, since the geometric areas of the electrodes are equal but wrinkled electrodes contain more surface area. With each aptamer containing one MB molecule, we correlate the higher MB signal with a greater amount of aptamers on the wrinkled electrodes. Normalizing the absolute peak height by the surface area calculated from integration of peak heights in sulfuric acid, we calculated the MB peak current density to be similar between standard and wrinkled electrodes (Figure 3B).

***Selectivity tests on individual SDW electrodes and S1 protein titration curve in 10% saliva:***

**(A)**

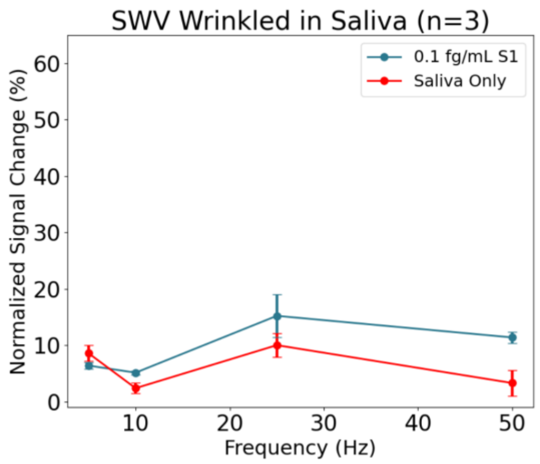


**(B)**

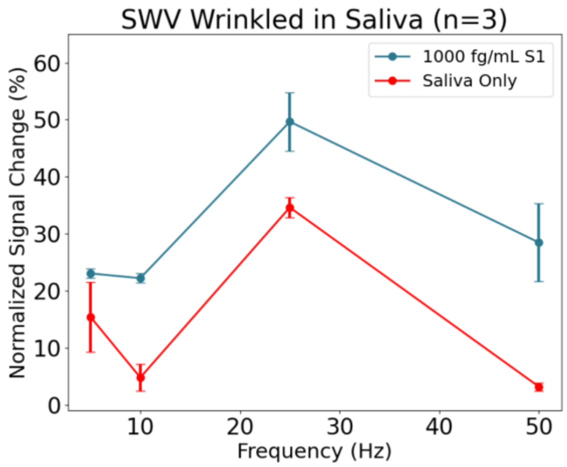


**(C)**

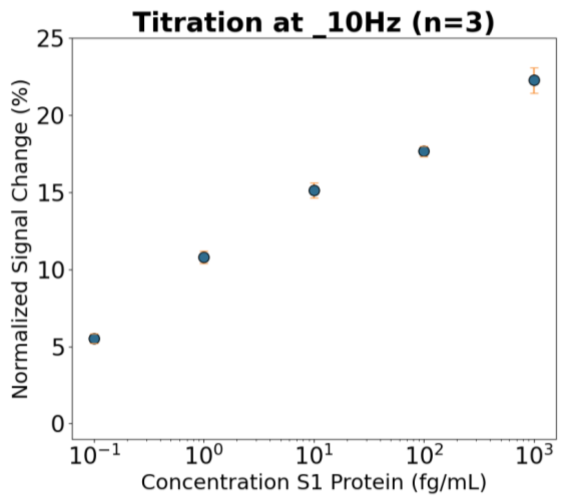


**(D)**

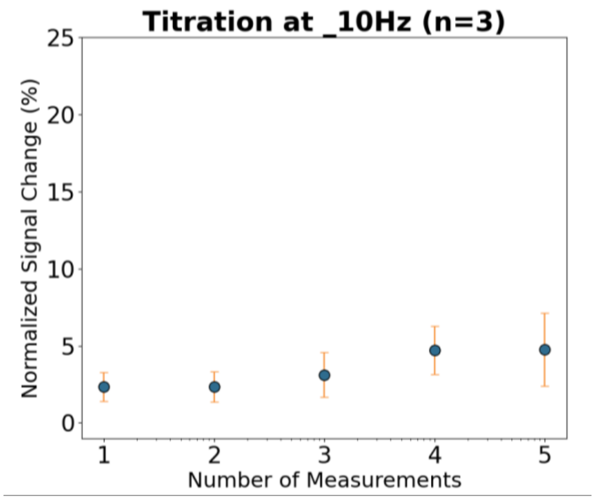


**Figure S2.** Measurements collected at all frequencies comparing signal change magnitude between individual SDW electrodes exposed to saliva and SDW electrodes exposed to saliva spiked with (A) 0.1 fg/mL S1 protein and (B) 1000 fg/mL S1 protein, which represent the range of the concentrations used in the titration curve. (C) Change in signal at 10 Hz with sequential addition of increasing concentrations of spike protein in 10% saliva. (D) Change in signal at 10 Hz with sequential addition of 10% saliva without spike protein.

SDW electrodes were found to be selective at two frequencies through the entire range of tested concentrations of S1 protein: 10 and 50 Hz. The data for spike and non-spiked saliva were extracted from the titration curve experiment shown in Figure 3, during which three electrodes were incubated multiple times with non-spiked saliva while a separate set of three electrodes were incubated with increasing concentrations of S1 protein in saliva. The times of incubation were equal (1 hr) between the non-spiked and spiked saliva.

The last incubation of non-spiked saliva was thus compared to the last concentration of S1 (1000 fg/mL). This is why the change in signal from non-spiked saliva in Figure S2 (B) is higher compared to Figure S2 (A). The electrodes had all been incubated several times with non-spiked saliva, to which we owe the greater signal change from saliva (compared to the first measurement).

***SDW mini-cell electrochemical characterization:***

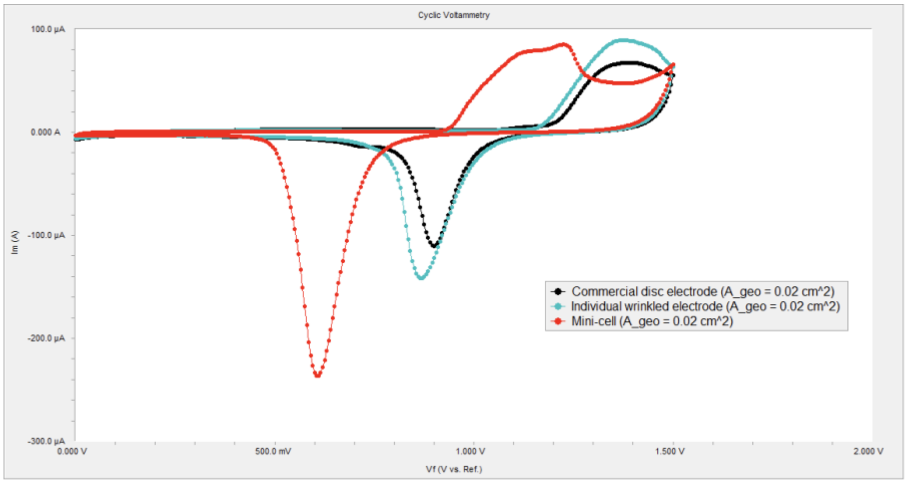


**Figure S3.** Comparison of commercial Au disc electrode, individual wrinkled electrode, and mini-cell. All electrodes have equivalent geometric areas. Measurements collected in 0.5 M sulfuric acid at 1000 mV/s.

***SDW electrode functionalization and stability assessment:***

Electrode functionalization and passivation time was determined for wrinkled electrodes in the following fashion. From the success of standard electrodes, the same protocol was first applied to wrinkled electrodes with minimal success. The incubation of  MCH was first adjusted from 18 hr (not shown) to 24 hr and then to 39 hr (Figure S2A and Figure S2B). This resulted in a marginally more stable signal. Following this, the aptamer incubation step, which precedes the MCH step, was extended from the original 4 hr to 8 hr (Figure S2C and Figure S2D). This resulted in a significant improvement to signal stability- defined here as the change in signal across 10 cycles of square wave voltammetry. If a sensor was found to have a baseline drift across 10 measurements that exceeded 5% normalized change, it was discarded and not used in experiments due to unreliable signal change reporting.

(A) 4 hr aptamers, 24 hr MCH

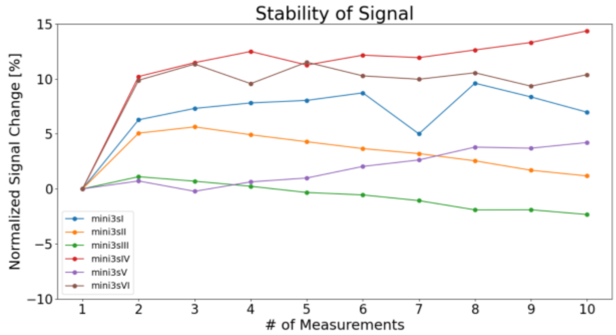


(B) 4 hr aptamers, 39 hr MCH

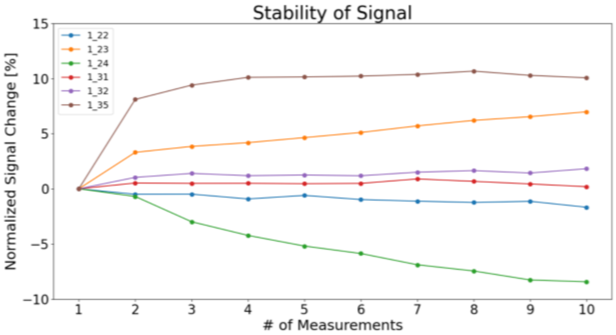


(C) 8 hr aptamers, 39 hr MCH

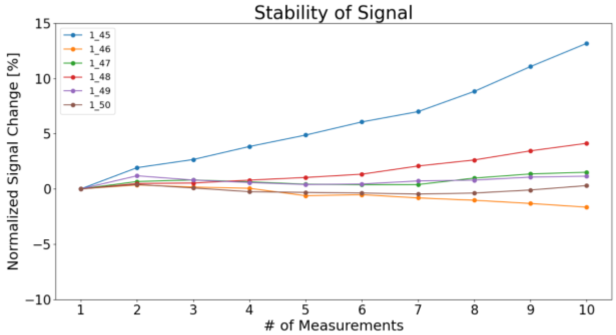


**Figure S4.** Signal stability was measured at 10 Hz to evaluate the change in signal across time prior to performing titration curves. Signal stability was compared between protocols with 4 hr aptamer incubation steps and (A) a 24 hr MCH incubation step and (B) a 39 hr MCH incubation step. Signal stability was then compared to (C) an 8 hr aptamer incubation steps measured after a 39 hr MCH incubation.

***Calculation of SDW mini-cell system noise[1] at 10 Hz:***

The system noise was inferred by dividing the standard deviation at or near zero concentration (0.1 fg/mL) by the square root of the number of samples then multiplying the quotient by 3.

1.89 (Standard deviation of normalized % change in signal at 0.1 fg/mL) / √3 (number of samples) * 3 = **3.29%** (normalized change in signal at the level of system noise)

This value is less than that measured upon introduction of the first concentration of S1 protein in saliva introduced to the SDW mini-cells, and we conclude that our sensor is sensitive to 0.1 fg/mL S1 protein in 10% saliva.

***References***

1 The fitness for purpose of analytical methods: a laboratory guide to method validation and related topics, 2014.
